# Thyroid function and the risk of Alzheimer’s disease: A Mendelian Randomisation study

**DOI:** 10.1101/2021.06.20.21259140

**Authors:** Eirini Marouli, Lina Yusuf, Alisa D. Kjaergaard, Rafat Omar, Aleksander Kuś, Oladapo Babajide, Rosalie Sterenborg, Bjørn O. Åsvold, Stephen Burgess, Christina Ellervik, Alexander Teumer, Marco Medici, Panos Deloukas

## Abstract

**Background:** Observational epidemiological studies propose that variations in thyroid function are associated with the risk of dementia; however, there have been inconsistent findings regarding the direction of the association. It is still unclear whether these associations are causal or not. We aim to test whether genetically determined variation within the normal range of thyroid function and hypothyroidism are causally associated with the risk of Alzheimer’s disease (AD).

**Methods:** Mendelian Randomisation (MR) analyses_using genetic instruments associated with normal range TSH and FT4 levels. Secondary analyses included investigation of the role of hypothyroidism. Bidirectional MR was conducted to address the presence of a potential reverse causal association. Summary statistics were obtained from the ThyroidOmics Consortium and the latest available GWAS meta-analyses.

**Results:** MR analyses show a causal association between normal range TSH levels and decreased risk of AD (OR = 0.98, 95% CI=0.97-0.99, p = 0.01). There was no evidence for a causal association between FT4 or hypothyroidism and Alzheimer’s disease. Bidirectional MR did not show any effect of Alzheimer’s disease on TSH or FT4.

**Conclusions:** This Mendelian Randomisation study shows for the first time a causal association between normal-range thyroid function and AD. Future studies should clarify the underlying pathophysiological mechanisms.

## Introduction

Dementia is an umbrella term comprising several neurodegenerative brain disorders which are characterised by the deterioration of cognitive functions including memory, planning, language, recognizing and reasoning. Alzheimer’s disease, Frontotemporal dementia, Lewy body dementia and Vascular dementia are four of the most common dementias (Gale, Acar, & Daffner, 2018). AD is the most common form of dementia, which is the fifth leading cause of death. AD can hinder the ability of an individual to perform everyday tasks along with several aspects of daily life. Despite decades of research only a few causal risk factors have been identified, and those which have been identified are mostly non-modifiable.

Levels of thyroid hormones are thought to impact the risk of developing dementia. Thyroid hormones are known for their involvement in brain development; they influence adult neurogenesis (Kapoor, Fanibunda, Desouza, Guha, & Vaidya, 2015). Many studies have investigated the association between thyroid function and dementia. The Rotterdam study (Chaker et al., 2016) found that higher TSH levels are linked to a lower risk of developing dementia, both within the full and normal ranges of thyroid function. This same study also found that high free thyroxine (FT4) is associated with an increased dementia risk. However, their MRI findings reveal that thyroid function is not linked to subclinical vascular brain disease, therefore, they concluded that high and high-normal thyroid function is associated with a greater dementia risk, but through non-vascular pathways.

A meta-analysis was carried out which included a total of 11 studies that were published between 2003 and 2016: three case-control studies and eight cohort studies. These were used to examine the relationship between FT4 and TSH with dementia. The results found that higher levels of FT4 were associated with an increased dementia risk. The meta-analysis findings also revealed that lower TSH levels were linked to an increased dementia risk, more specifically, the link was confined to TSH levels below the normal range, rather than levels within the normal range (Wu, Pei, Wang, Xu, & Cui, 2016).

Conversely, the Framingham study conducted by (Tan et al., 2008) examined the relationship between thyroid function and Alzheimer’s disease and found that both high and low TSH levels within the normal range were associated with a more than 2-fold greater Alzheimer’s disease risk in women but found no association between TSH and AD in men. Another study by (van Osch, Hogervorst, Combrinck, & Smith, 2004) also found that lower TSH levels within the reference range were associated with an increased AD risk; however, in contrast to the Framingham study, these results were independent of gender.

At present, most studies that have examined the relationship between thyroid function and dementia have been cross-sectional. In addition, their sample sizes have been small, compared to large scale meta-analyses available now, which has been a limitation and reduces power. There have been inconsistent findings regarding the nature of the thyroid function and dementia relationship; for example, some large prospective studies have found that elevated levels of FT4 are associated with an increased dementia risk (de Jong et al., 2009; Wu et al., 2016), whilst other studies have found no association (Gussekloo et al., 2004). It is not clear whether variation in thyroid function results from or contributes to Alzheimer’s disease and whether the underlying associations are causal or not.

Mendelian Randomisation (MR) is an observational epidemiological method which uses genetic data, normally in the form of single nucleotide polymorphisms (SNPs), to investigate causal relationships between health outcomes and associated risk factors (Neil M. D., 2018).

One benefit of using MR is its ability to overcome reverse causality and confounding; which is when a factor affects both the health outcome and associated risk factor, this can lead to biased results (Zheng et al., 2017). Mendel’s Laws of Inheritance states that alleles segregate randomly into gametes; the genetic variants used for MR analysis are assorted during the formation of gametes, therefore are not confounded by lifestyle choices or environmental factors (Zheng et al., 2017). We currently have an important number of genetic instruments for thyroid function and it is now the optimal moment to perform MR analyses and evaluate the role of thyroid function on AD.

The aim of this manuscript is to test whether genetically determined variation within the normal range of thyroid function is associated with the risk of Alzheimer’s disease.

## Materials and methods

Main analyses examined whether there was any causal association between variation in normal range thyroid function evaluated by TSH and FT4 levels, and the risk of Alzheimer’s disease.

Secondary analyses examined whether hypothyroidism is causally associated with the risk of developing Alzheimer’s disease. Sensitivity analyses evaluated the role of thyroid function with estimates derived from GWAS limited in individuals with TSH within the normal range, aged<50 years old or the full range of TSH.

### Genetic variants used as instruments

We used 86 and 29 genetic variants respectively that were found to be associated at a genome-wide significant level (p<5×10^−8^), with TSH ((Zhou et al., 2020) and FT4 (Teumer et al, 2018). We also evaluated the effect of 93 genetic variants associated with hypothyroidism (Saevarsdottir et al., 2020). For TSH we also used estimates from the GWAS meta-analysis in the HUNT study for full range TSH levels, TSH within the normal range and using data only from individuals <50 years old (Zhou et al., 2020).

### Summary statistics and study population

The datasets on thyroid function derived from the ThyroidOmics Consortium (Teumer, 2018) and (Zhou et al., 2020), which carried out the largest meta-analysis on thyroid function. Secondary analyses for TSH were performed using estimates from GWAS data for full range TSH levels (HUNT + MGI + ThyroidOmics meta-analysis), TSH within the normal range (HUNT study) and TSH levels in individuals younger than 50 years old (HUNT study) (Zhou et al., 2020).

The summary statistics for Alzheimer’s disease were obtained from the GWAS Catalog involving a meta-analysis including 71,880 cases and 383,378 controls (Jansen et al., 2019).

### Two-sample Mendelian Randomisation

Two-sample MR was performed using the genetic data that were extracted from the GWAS summary statistics. We employed the Two-sample MR package in R (Hemani et al., 2018). We used the Inverse Variance Weighted (IVW) estimator (random effects), which assumes that there is no pleiotropy and is the most optimal as it maximises the likelihood function and achieves minimal variance (Lee, Cook, Lee, & Han, 2016). For variants that are not in linkage disequilibrium, the Inverse Variance Weighted estimate can be obtained from an IVW meta-analysis of the individual variants’ ratio estimates (S Burgess et al., 2020).

Sensitivity analyses were also performed using methods such as Mendelian Randomisation-Egger (MR-Egger). The MR-Egger method is a robust statistical technique that allows all the variants to have pleiotropic effect on the condition that these effects are not proportional to the effect that the variant has on the risk factor of interest; this is known as the Instrument Strength Independent of Direct Effect (InSIDE) assumption (Bowden, Davey Smith, & Burgess, 2015). This approach provides less precise estimates than the IVW and weighted median methods, thus, lowering the power for testing causal hypotheses (Neil M. D., 2018). However, it still gives consistent causal effect estimates regardless of the possibility that all the genetic variants may be invalid instrumental variables. During this investigation, a pleiotropy test was carried out, to evaluate the Egger intercept. This is an indicator to evaluate directional horizontal pleiotropy (Hemani et al., 2018).

### MR-PRESSO

The Mendelian Randomisation Pleiotropy RESidual Sum and Outlier (MR-PRESSO) approach was also used to evaluate the role of pleiotropy. When testing the causal null hypothesis, biased causal estimates and increased type I error rates are often the outcome of having the presence of pleiotropic variants in the MR study (S. Burgess & Thompson, 2013). The first test carried out was the ‘MR-PRESSO global test’ which evaluates the overall horizontal pleiotropy. The test compares the observed distance of the variants to the regression line with the expected distance (Verbanck, Chen, Neale, & Do, 2018). The second test carried out was the ‘MR-PRESSO outlier test’ which identifies variants that could be leading to horizontal pleiotropy and are subsequently removed. This test used the observed and expected distributions of the tested variant to identify the outlier variants. The third test carried out was the ‘MR-PRESSO distortion test’ which tested for significant differences in the causal estimates pre- and post-removal of the horizontal pleiotropic variants (Verbanck et al., 2018)

### Bidirectional MR

Bidirectional Mendelian Randomisation was carried out to examine the potential of a reverse causation (Sekula, Del Greco, Pattaro, & Köttgen, 2016). This approach investigated whether Alzheimer’s disease could have a causal effect on thyroid function. The MR-PRESSO method was also used to test for horizontal pleiotropy. We performed LD pruning for the genome-wide AD associated variants used as instruments (Jansen I et al, 2019).

## Results

### Normal range TSH and FT4 levels

MR analyses suggest that genetically determined TSH is associated with Alzheimer’s disease. (OR = 0.98, 95% CI=0.97-0.99, p = 0.01, IVW, per 1 Standard Deviation (SD) increase in TSH levels). In the MR PRESSO analysis, The Global Test returned a significant p-value indicating that overall horizontal pleiotropy was identified. Next, we identified these pleiotropic variants using the outlier test. The MR estimate remained significant (p=0.008) after removal of these horizontal pleiotropic outliers (Figure 1).

**Figure 1.**
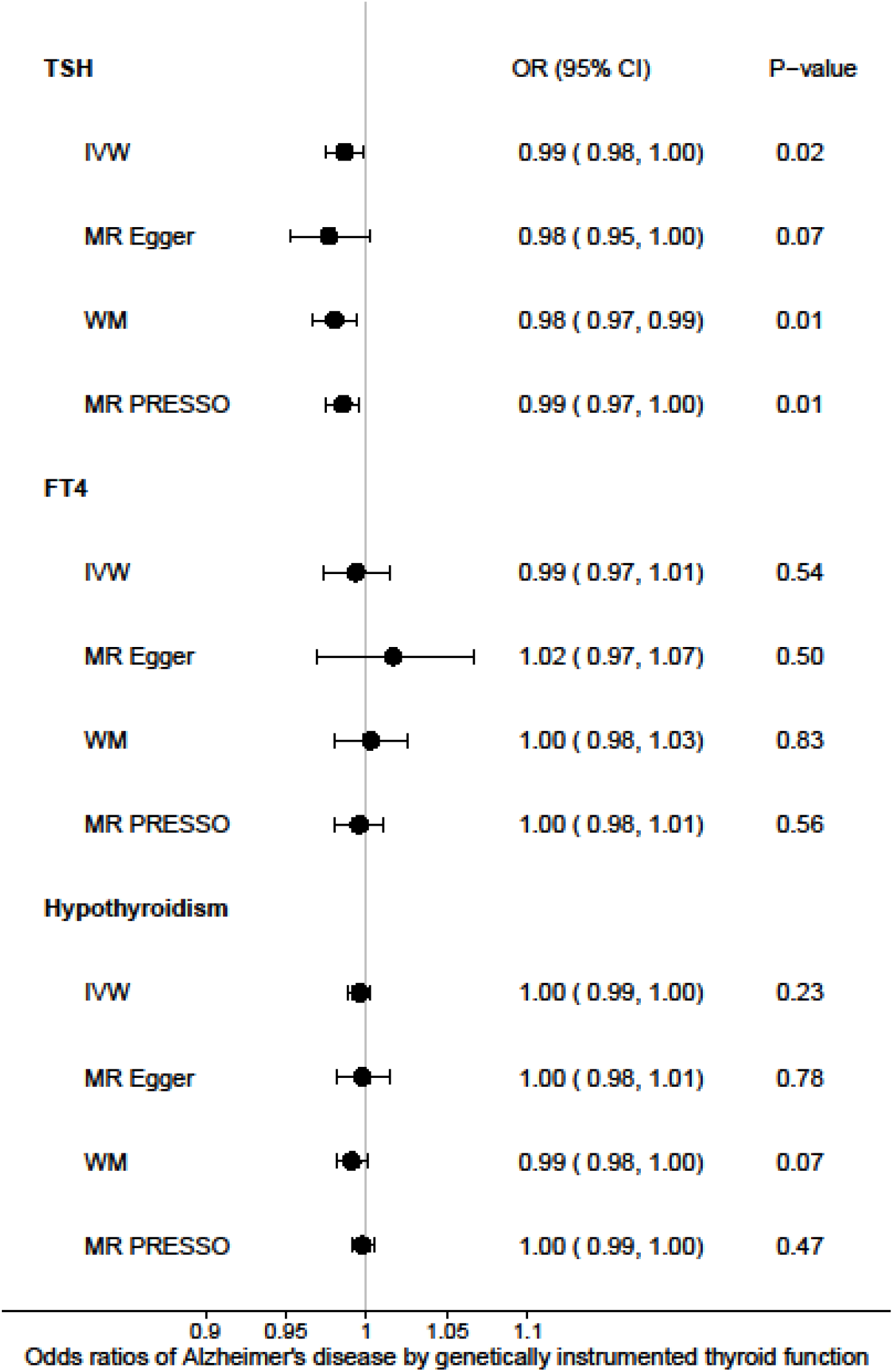
Forest plot = Odds ratios for the effect of genetically determined TSH, FT4 and hypothyroidism on Alzheimer’s disease risk.

There was no evidence for causal association between normal range FT4 levels and the risk of developing Alzheimer’s disease (p = 0.49, MR-PRESSO). The Global Test returned a significant p-value indicating that overall horizontal pleiotropy was identified; the outlier test identified the pleiotropic variants and subsequently removed them. However, after correcting the horizontal pleiotropic outliers, the MR estimate remained null (p=0.56). (Figure 1)

### Secondary analyses

Secondary analyses using estimates from GWAS data for full range TSH levels (HUNT + MGI + ThyroidOmics), within the normal range (HUNT study) and TSH levels for individuals younger than 50 years old (HUNT study), revealed nominally significant associations with Alzheimer’s disease. More specifically: full-range TSH: p = 0.06, TSH within normal range: p=0.04 and participants younger than 50 years old: p = 0.04 (Figure 2). MR-PRESSO Global Test P-value was significant for all 4 sets indicating the detection of overall horizontal pleiotropy. However, after the removal of horizontal pleiotropic outliers, the MR estimates remained non-significant (Supplementary Tables). There was no evidence for the causal association between hypothyroidism and Alzheimer’s disease (Hypothyroidism: p= 0.99).

**Figure 2.**
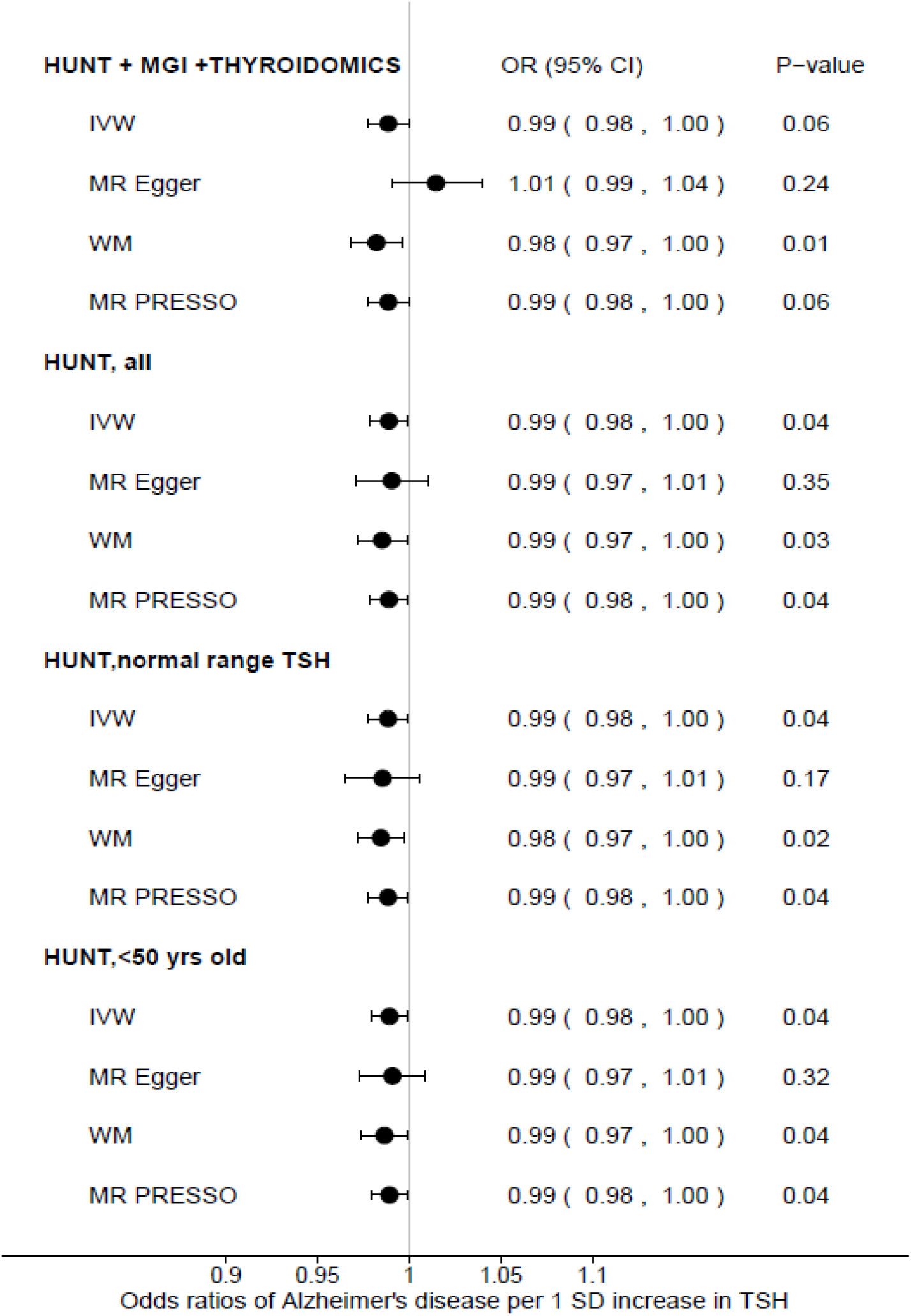
Forest plot for the effect of genetically determined TSH on Alzheimer’s disease risk using summary statistics for full range TSH levels (HUNT + MGI + ThyroidOmics meta-analysis), full range TSH levels (HUNT study), TSH within the normal rang in the HUNT study, and TSH levels in individuals younger than 50 years old (HUNT study)

### Bidirectional MR analyses

Bidirectional MR was carried out to address a potential reverse causal association and investigate whether Alzheimer’s disease has a causal effect on TSH levels. Across all MR methods, the results suggest no evidence for causal association between Alzheimer’s disease and normal range TSH of FT4 levels. In the MR PRESSO analysis, The Global Test returned a non-significant p-value suggesting no evidence for horizontal pleiotropy.

## Discussion

This study’s main aim was to explore the causal relationship between thyroid function and Alzheimer’s disease with the use of Mendelian Randomisation techniques. We show for the first time that increased levels of genetically determined TSH within the normal range were significantly associated with a decreased risk of AD. This finding is of particular importance as thyroid function is a modifiable risk factor. Both normal range FT4 levels and hypothyroidism had no significant associations with Alzheimer’s disease. Bidirectional analyses showed no causal effect of the genetically determined Alzheimer’s disease on TSH or FT4.

There are several inconsistencies in previous observational studies regarding the relationship between thyroid function and AD. A recent study that looked at the relationship between decreased regional cerebral blood flow (rCBF), associated with Alzheimer’s, and thyroid function found that there were significant correlations between TSH and Rcbf (Nomoto et al., 2019). However, this result was only seen within patients exhibiting mild cognitive impairment and not in patients with AD. The study also found that the correlation between FT3 and rCBF was significant in AD. The Framingham study of 2008 found that both the upper and lower tertiles of TSH in the normal range indicated an increased risk of dementia in women (Tan et al., 2008), but not in men. Wu et al’s meta-analysis showed that high FT4 and low TSH outside the normal range were significantly associated with AD (Wu et al., 2016). These findings partly concur with our findings as low TSH levels within the normal range were found to be a risk factor for AD.

Tan et al theorised that the link between thyroid function and Alzheimer’s disease is the aggregation of amyloid β peptides which leads to less Thyrotropin-releasing hormone (TRH) being secreted by the hypothalamus. This affects the TRH’s role within the pituitary gland, thus decreasing the production of TSH leading to a decrease of thyroxine levels (Tan & Vasan, 2009). This directly opposes the meta-analysis study which found that higher FT4 and lower TSH was a risk factor in patients developing Alzheimer’s (Wu et al., 2016).

The theory also implies that AD had a causal relationship on TSH and FT4, rather than the inverse. Our study’s bidirectional analysis did not show any significant causal relationship between AD and thyroid function. A potential cause for the differing results when looking at the relationship between AD and thyroid function could be that some studies were cross-sectional looking at the correlations as opposed to the causal relationship.

This is a well-powered study that could detect even small effects of variation within normal-range thyroid function on Alzheimer’s disease. There are many strengths for using MR as a method to find causal relations and capable of reducing confounding. The summary statistics used were from the largest GWAS published to increase statistical power. Pleiotropy was tested using sensitivity analyses, and possible pleiotropic variants were removed, which did not significantly change the results. The use of MR approaches to investigate non-confounded causal associations is becoming more prevalent due to the many advantages that the method has over more traditional observational epidemiological studies. Furthermore, multiple methods of MR were used to reduce biased estimates which helped in finding consistent results. A potential limitation of our study is that we focused our analyses on individuals of European ancestry, suggesting that these results could not be easily transferred to other populations. In addition, we potentially did not observe a signal between hypothyroidism and AD, due to the fact that various autoimmune diseases congregate in hypothyroidism definition and we therefore used summary-statistics from a quite heterogeneous group of instruments.

## Conclusion

We show that increased levels of genetically determined TSH within the normal range are associated with a decreased risk of Alzheimer’s disease. In contrast, our results do not show any evidence for a causal association between neither FT4 nor hypothyroidism with AD. These results are clinically relevant; as appropriate management of thyroid function could improve memory. These results have a key role in guiding clinicians towards thyroid function optimization in Alzheimer’s disease patients. Our work could pave the way towards further studies involving randomized control trials for thyroid function and Alzheimer’s disease and further studies are needed to clarify the exact underlying mechanism.

## Data Availability

All data information is provided in the manuscript.

## Abbreviations

FT4: Free thyroxine
IVW: Inverse variance weighted
LD: Linkage disequilibrium
MR: Mendelian Randomisation
OR: Odds ratio
SD: Standard deviation
SNP: Single nucleotide polymorphism
TSH: Thyroid stimulating hormone

## Notes

### Competing Interest Statement

The authors have declared no competing interest.

### Clinical Trial

NA

### Funding Statement

The Barts Biomedical Research Centre funded by the UK National Institute for Health Research (NIHR) has supported COVID related research.

## References

Bowden, J., Davey Smith, G., & Burgess, S. (2015). Mendelian randomization with invalid instruments: effect estimation and bias detection through Egger regression. Int J Epidemiol, 44(2), 512–525. doi:10.1093/ije/dyv080

Burgess, S., Davey Smith, G., Davies, N., Dudbridge, F., Gill, D., Glymour, M.,... Theodoratou, E. (2020). Guidelines for performing Mendelian randomization investigations [version 2; peer review: 2 approved]. Wellcome Open Research, 4(186). doi:10.12688/wellcomeopenres.15555.2

Burgess, S., & Thompson, S. G. (2013). Use of allele scores as instrumental variables for Mendelian randomization. Int J Epidemiol, 42(4), 1134–1144. doi:10.1093/ije/dyt093

Chaker, L., Wolters, F. J., Bos, D., Korevaar, T. I., Hofman, A., van der Lugt, A.,... Ikram, M. A. (2016). Thyroid function and the risk of dementia: The Rotterdam Study. Neurology, 87(16), 1688– 1695. doi:10.1212/wnl.0000000000003227

de Jong, F. J., Masaki, K., Chen, H.,... Remaley, A. T., Breteler, M. M., Petrovitch, H., Launer, L. J. (2009). Thyroid function, the risk of dementia and neuropathologic changes: the Honolulu-Asia aging study. Neurobiol Aging, 30(4), 600–606. doi:10.1016/j.neurobiolaging.2007.07.019

Gale, S. A., Acar, D., & Daffner, K. R. (2018). Dementia. The American Journal of Medicine, 131(10), 1161–1169. doi:10.1016/j.amjmed.2018.01.022

Gussekloo, J., van Exel, E., de Craen, A. J., Meinders, A. E., Frölich, M., & Westendorp, R. G. (2004). Thyroid status, disability and cognitive function, and survival in old age. Jama, 292(21), 2591– 2599. doi:10.1001/jama.292.21.2591

Hemani, G., Zheng, J., Elsworth, B., Wade, K. H., Haberland, V., Baird, D.,... Haycock, P. C. (2018). The MR-Base platform supports systematic causal inference across the human phenome. eLife, 7, e34408. doi:10.7554/eLife.34408

Jansen, I. E., Savage, J. E., Watanabe, K., Bryois, J., Williams, D. M., Steinberg, S.,... Posthuma, D. (2019). Genome-wide meta-analysis identifies new loci and functional pathways influencing Alzheimer’s disease risk. Nature Genetics, 51(3), 404–413. doi:10.1038/s41588-018-0311-9

Kapoor, R., Fanibunda, S. E., Desouza, L. A., Guha, S. K., & Vaidya, V. A. (2015). Perspectives on thyroid hormone action in adult neurogenesis. Journal of Neurochemistry, 133(5), 599–616. doi:https://doi.org/10.1111/jnc.13093

Lee, C. H., Cook, S., Lee, J. S., & Han, B. (2016). Comparison of Two Meta-Analysis Methods: Inverse-Variance-Weighted Average and Weighted Sum of Z-Scores. Genomics & informatics, 14(4), 173–180. doi:10.5808/GI.2016.14.4.173

Neil M. D. M. V. H., George D. S. (2018). Reading Mendelian randomisation studies: a guide, glossary, and checklist for clinicians. British Medical Journal 362.

Nomoto, S., Kinno, R., Ochiai, H., Kubota, S., Mori, Y., Futamura, A.,... Ono, K. (2019). The relationship between thyroid function and cerebral blood flow in mild cognitive impairment and Alzheimer’s disease. PloS one, 14(4), e0214676–e0214676. doi:10.1371/journal.pone.0214676

Saevarsdottir, S., Olafsdottir, T. A., Ivarsdottir, E. V., Halldorsson, G. H., Gunnarsdottir, K., Sigurdsson, A., Stefansson, K. (2020). FLT3 stop mutation increases FLT3 ligand level and risk of autoimmune thyroid disease. Nature, 584(7822), 619–623. doi:10.1038/s41586-020-2436-0

Sekula, P., Del Greco, M. F., Pattaro, C., & Köttgen, A. (2016). Mendelian Randomization as an Approach to Assess Causality Using Observational Data. (1533-3450 (Electronic)).

Tan, Z. S., Beiser, A., Vasan, R. S., Au, R., Auerbach, S., Kiel, D. P.,... Seshadri, S. (2008). Thyroid function and the risk of Alzheimer disease: the Framingham Study. Archives of internal medicine, 168(14), 1514–1520. doi:10.1001/archinte.168.14.1514

Tan, Z. S., & Vasan, R. S. (2009). Thyroid Function and Alzheimer’s Disease. Journal of Alzheimer’s Disease, 16, 503–507. doi:10.3233/JAD-2009-0991

Teumer, A. (2018). Common Methods for Performing Mendelian Randomization. Frontiers in Cardiovascular Medicine, 5(51). doi:10.3389/fcvm.2018.00051

van Osch, L. A., Hogervorst, E., Combrinck, M., & Smith, A. D. (2004). Low thyroid-stimulating hormone as an independent risk factor for Alzheimer disease. Neurology, 62(11), 1967–1971. doi:10.1212/01.wnl.0000128134.84230.9f

Verbanck, M., Chen, C.-Y., Neale, B., & Do, R. (2018). Detection of widespread horizontal pleiotropy in causal relationships inferred from Mendelian randomization between complex traits and diseases. Nature Genetics, 50(5), 693–698. doi:10.1038/s41588-018-0099-7

Wu, Y., Pei, Y., Wang, F., Xu, D., & Cui, W. (2016). Higher FT4 or TSH below the normal range are associated with increased risk of dementia: a meta-analysis of 11 studies. Sci Rep, 6, 31975. doi:10.1038/srep31975

Zheng, J., Baird, D., Borges, M.-C., Bowden, J., Hemani, G., Haycock, P.,... Smith, G. D. (2017). Recent Developments in Mendelian Randomization Studies. Current Epidemiology Reports, 4(4), 330–345. doi:10.1007/s40471-017-0128-6

Zhou, W., Brumpton, B., Kabil, O., Gudmundsson, J., Thorleifsson, G., Weinstock, J.,... Åsvold, B. O. (2020). GWAS of thyroid stimulating hormone highlights pleiotropic effects and inverse association with thyroid cancer. Nature Communications, 11(1), 3981. doi:10.1038/s41467-020-17718-z

